# “The more I fear about COVID-19, the more I wear medical masks”: A survey on risk perception and medical masks’ uses

**DOI:** 10.1101/2020.03.26.20044388

**Authors:** Toan Luu Duc Huynh

## Abstract

The legal behaviors in using medical masks in public have been finally promulgated by the Vietnamese Government after 47 days since the WHO declared the Public Health Emergency of International Concern (PHEIC) due to the COVID-19 pandemic. From a sample of 345 Vietnamese respondents aged from 15 to 47 years, this brief note found that the risk perception of COVID-19 danger significantly increases the likelihood of wearing the medical masks. In addition, there is a weak evidence about the differences in age under the COVID-19 outbreaks. More noticeably, those who use masks before COVID-19 pandemic tend to maintain their behaviors. Our results offer the insightful into Vietnamese citizens’ responses in terms of using medical masks; even the uses of this method are still controversial. Our results are robust by performing Exploratory Factor Analysis for five features and further regressions.

## 1. Introduction

Obviously, humankind is currently facing a global health crisis. Our lives experience uncertainty about the massive scale of the epidemic due to the easy transmission of the second type of coronavirus, which is known as the COVID-19 pandemic as the Public Health Emergency of International Concern by World Health Organization. The decisions people and government take in the previous days might not only shape society for years but also create new social norms or cultural norms. Many existing studies have raised the concern about policy intervention (Haushofer & Metcalf, 2020), suspending the educational system (Zhang et al., 2020), risk management in universities (Wang et al., 2020), seven behaviors’ intervention (Lunn et al, 2020) etc. However, there are growing bodies of examining the impact of COVID-19 outbreaks on different socioeconomic and political aspects in both a single country and the global scope.

Vietnam is known as the first country to succeed in the local containment of SARS in 2003 (Cao et al., 2019). In addition, this is one of the countries which limited local transportation (Gilbert et al., 2020). More importantly, Vietnam has the successfully beginning step to in containment of COVID-19 when closing the border as well as organizing the compulsory quarantine at the military areas for controlling strictly. There are many policies implemented since the first case of COVID-19 was found in Vietnam. For instance, Vietnam stopped issuing new visas to all foreign nationals for 30 days in a bid to curb the spread of COVID-19. In February 2020, the Vietnamese Ministry of Education and Training suspended all school activities across the country until the end of March as part of quarantine measures against the spreading of the virus. It later extended this till the middle of April until further notice. More noticeably, on 16 March 2020, the Vietnamese government requested that everyone wear face masks when going to public areas to protect themselves and others.

Although the World Health Organization recommends that only positively-confirmed patients should wear medical masks to mitigate COVID-19 transmission, Vietnam is the first country to legalize the behaviors of using medical masks in public. It is not an optional choice anymore. Since this regulation becomes effective, the Vietnamese citizens have to self-protect by wearing medical masks in the public area. However, the use of medical masks as one of the prevention methods is still debatable. For example, Leung et al. (2020) suggested that there should have guidance for mass masking. However, WHO recommends that there exists a lack of evidence (WHO, 2020). Up to date, health authorities suggest social distancing, hand-washing, and so forth.

This short communication focuses on risk perception and medical masks wearing of Vietnamese citizens by conducting the voluntary survey for 345 Vietnamese respondents aged from 15 to 47 years. Although we do find several results regarding the use of medical masks in Vietnam, come caveats are essential. Our study has been analyzed much more rapidly from preliminary evaluation than would be standard for research of this type.

In doing so, we contribute to the existing literature by four main points. Firstly, the risk perception of COVID-19 danger significantly increases the likelihood of wearing medical masks. Secondly, we find that before the COVID-19 epidemic, the elder is not likely to wear medical masks. However, after the COVID-19 outbreaks, the rise in age significantly increases the likelihood of using masks. Thirdly, price is the only factor that significantly decreases the willingness to buy medical masks. Fourthly and finally, those who use masks before the COVID-19 pandemic tend to maintain their behaviors. We are running to fight the virus, and the COVID-19 study has to trade thoroughness off against speed, at least to some extent. However, based on our dataset, our analytical results are striven to prioritize the accuracy as well as potential policymakers.

The short paper is organized as follows. Section 2 describes the data collection. We analyze and report our findings in Section 3. Section 4 concludes.

## 2. Data collection and descriptive statistics

### 2.1. Questionnaire design

The questionnaire consisted of three parts. The first section included the socioeconomics questions about age, gender, family size, and religion. The second part had the risk perception questions as well as medical masks’ behaviors (the use before the outbreak, the binary choice, the number of medical mask boxes). Moreover, the five characteristics of medical masks (price, brand, function, origin, and safeness) are also a part of our questionnaire. The questionnaire was mainly based on the Vietnamese language. Data were collected since February 1, 2020, when the Vietnamese prime minister officially declared the global and national emergency scenario. The duration of the survey was 20 working days, representing three weeks after the announcement of the epidemic. The average time to fulfill the questionnaire is 10 minutes. The questionnaire is randomly circulated by the respondents without any intervention from the researchers. As mentioned earlier, this survey is open for 20 working days; therefore, the platform is automatically closed on the expired date. In addition, this survey is approved by the President of Scientific Committee in School of Banking, University of Economics Ho Chi Minh City about the ethics declaration. The data were analyzed anonymously.

### 2.2. Descriptive statistics of the sample

A total of 395 respondents were recruited from randomized sources: Internet online platform. In most cases, the questionnaire is randomly distributed within 20 working days. Of the total 395 participants answering the questionnaire, 50 were omitted due to too few main answers in part 2 or having the ineligible answers (for example, they answered that they don’t want to buy the medical masks in binary choice; however they estimated the number of medical mask boxes). Therefore, we excluded these respondents, and finally, our sample covers 345 observations for analyzing.

Table 1 summarizes demographic, socioeconomic characteristics and decision-making in medical masks choices. The age of our respondents ranged from 15 to 47 years and 92.75% of them fell into the age category of 18-35 years. Gender exhibits the balanced proportion in our sample with 45.58% men and 54.42% women. The majority of participants are non-religion (65.11%), Buddhism (30.49%) and other religion (4.40%).

**Table 1.** The data descriptive statistics and relevant questionnaire.

| Variable | Questions | Mean | Std. Dev. |
| --- | --- | --- | --- |
| AGE | How old are you? | 24.42 | 6.03 |
| FAMILY_SIZE | How many family members do you have? | 4.43 | 1.87 |
| RELIGION | Which religion are you belonging to? (0 - No religion, 1 - Buddhism, 2 - Others) | 0.39 | 0.57 |
| GENDER | What is your gender? (0 - Male, 1 - Female, 2 - Others) | 0.54 | 0.50 |
| BUY_MASK | At present, will you buy the medical masks to protect yourself against COVID-19? | 0.61 | 0.49 |
| USE_BEFORE | Before COVID-19, do you use medical mask at the public places? | 0.73 | 0.45 |
| RISK_PERCEPTION | How likely do you concern the COVID-19 pandemic? (1- least concern; 10 - most concern) | 7.65 | 1.80 |
| NUMBER_BOX | How many medical mask boxes would you like to buy in the next few days? | 50.84 | 72.13 |
| PRICE | To which extent do the price of medical masks applies to you to buy medical masks? (1- less concern; 10 - most concern) | 6.18 | 2.66 |
| BRAND | To which extent do the brand of medical masks applies to you to buy medical masks? (1- less concern; 10 - most concern) | 6.10 | 2.75 |
| FUNCTION | To which extent do the function of medical masks applies to you to buy medical masks? (1- less concern; 10 - most concern) | 7.34 | 2.67 |
| ORIGIN | To which extent do the origin of medical masks applies to you to buy medical masks? (1- less concern; 10 - most concern) | 7.07 | 2.74 |
| SAFENESS | To which extent do the safeness of medical masks applies to you to buy medical masks? (1- less concern; 10 - most concern) | 7.77 | 2.64 |

Overall, the Vietnamese citizen perceived the risk of COVID-19 outbreak as higher as the average value from 10-point scale (Mean = 7.65, S.D = 1.80, p < 0.001). In addition, it is worth noting that before the COVID-19 outbreak, 72.85% respondents choose to wear masks with the purpose of avoiding air-pollution and sun. However, only 61.46% participants in our sample chose to buy medical masks under the COVID-19 pandemic. From our sample, one respondent will choose to buy average 50.84 boxes (approximately 250 pieces in total). In addition, ‘*safeness*’ is the most concerning element (Mean = 7.77, S.D = 2.64) among five features of medical masks while people are not likely to care about the brand of medical mask (Mean = 6.10, S.D = 2.75).

## 3. Main findings

### 3.1. Differences in the medical mask use

Before the COVID-19 epidemic, there are only two factors such as age and risk perception having the mean differences between those who use medical masks and refuse to use. In particular, the average age of those who choose to use masks is 24.03 (S.D = 0.33) while the opposite group exhibits slightly higher (Mean = 25.36, S.D = 0.72), which has the weakly significant t-test with equal variances (p < 0.1). Additionally, there is a difference in risk perception between two groups before COVID-19 outbreak. To be more detail, those who use medical masks have the higher risk perception than the other (t-test, p < 0.001). It implies that people choose to wear medical mask before pandemic are ready to perceive more risk than the counterpart. We do not find any specific difference between two groups in terms of five masks characteristics. At present, we also asked the respondents whether they want to use medical masks as one of the protective methods to fight against the COVID-19 or not. Therefore, we would like to examine the differences in the medical mask use on two main categories: (i) socioeconomics, (ii) five mask features. Firstly, we find weak evidence in age difference between two groups (use and non-use of medical masks) (p < 0.1). Similarly, those who choose to buy medical masks have the higher risk perception than the other (t-test, p < 0.01). Lastly, there is no difference in five mask features between two groups.

To sum up, by using the simple t-test, we find that there is a strong evidence in risk perception between those who choose to wear (to buy) medical mask and the other. The finding in age group exhibits the weak statistics while the other determinants are not found.

### 3.2. Regression results

Before going to the main regression model, we also perform correlation test to examine whether multi-collinearity happens in our dataset or not. Table 2 represents the correlation among our variables. The socioeconomic factors have the lower correlated values while the mask features exhibit the strong correlation. Therefore, we are very cautious about multi-collinearity.

**Table 2.** Correlation matrix.

|  | AGE | FAMILYSIZE | RELIGION | GENDER | BUYMASK | USEBEFORE | RISKPERCEPTION | NUMBERBOX | PRICE | BRAND | FUNCTION | ORIGIN | SAFENESS |
| --- | --- | --- | --- | --- | --- | --- | --- | --- | --- | --- | --- | --- | --- |
| AGE | 1 |  |  |  |  |  |  |  |  |  |  |  |  |
| FAMILY_SIZE | -0.104 | 1 |  |  |  |  |  |  |  |  |  |  |  |
| RELIGION | 0.060 | 0.015 | 1 |  |  |  |  |  |  |  |  |  |  |
| GENDER | 0.001 | 0.030 | 0.016 | 1 |  |  |  |  |  |  |  |  |  |
| BUY_MASK | 0.094 | -0.046 | -0.065 | 0.070 | 1 |  |  |  |  |  |  |  |  |
| USE_BEFORE | -0.099 | -0.018 | 0.035 | 0.038 | 0.133 | 1 |  |  |  |  |  |  |  |
| ISK_PERCEPTION | 0.012 | -0.019 | 0.119 | -0.007 | 0.160* | 0.204* | 1 |  |  |  |  |  |  |
| NUMBER_BOX | 0.016 | 0.001 | -0.041 | 0.049 | 0.561* | 0.029 | 0.057 | 1 |  |  |  |  |  |
| PRICE | -0.146* | -0.063 | 0.068 | -0.027 | 0.039 | 0.083 | 0.138* | -0.050 | 1 |  |  |  |  |
| BRAND | -0.087 | -0.108 | 0.012 | -0.026 | 0.013 | 0.033 | 0.146* | 0.017 | 0.591* | 1 |  |  |  |
| FUNCTION | -0.152* | -0.013 | 0.028 | 0.005 | 0.052 | 0.017 | 0.117 | 0.051 | 0.712* | 0.735* | 1 |  |  |
| ORIGIN | -0.098 | -0.011 | -0.005 | 0.021 | 0.007 | 0.044 | 0.122 | 0.051 | 0.635* | 0.780* | 0.837* | 1 |  |
| SAFENESS | -0.112 | -0.052 | 0.024 | -0.024 | 0.023 | 0.009 | 0.063 | 0.023 | 0.695* | 0.683* | 0.910* | 0.849* | 1 |

Notes: \* denotes the significance level at 1%.

Table 3 demonstrates the regression results to predict the behaviors in using medical masks before and after the COVID-19 outbreaks. In addition, our regression results also emphasize the prediction to the number of medical mask boxes bought by Vietnamese citizens.

**Table 3.**
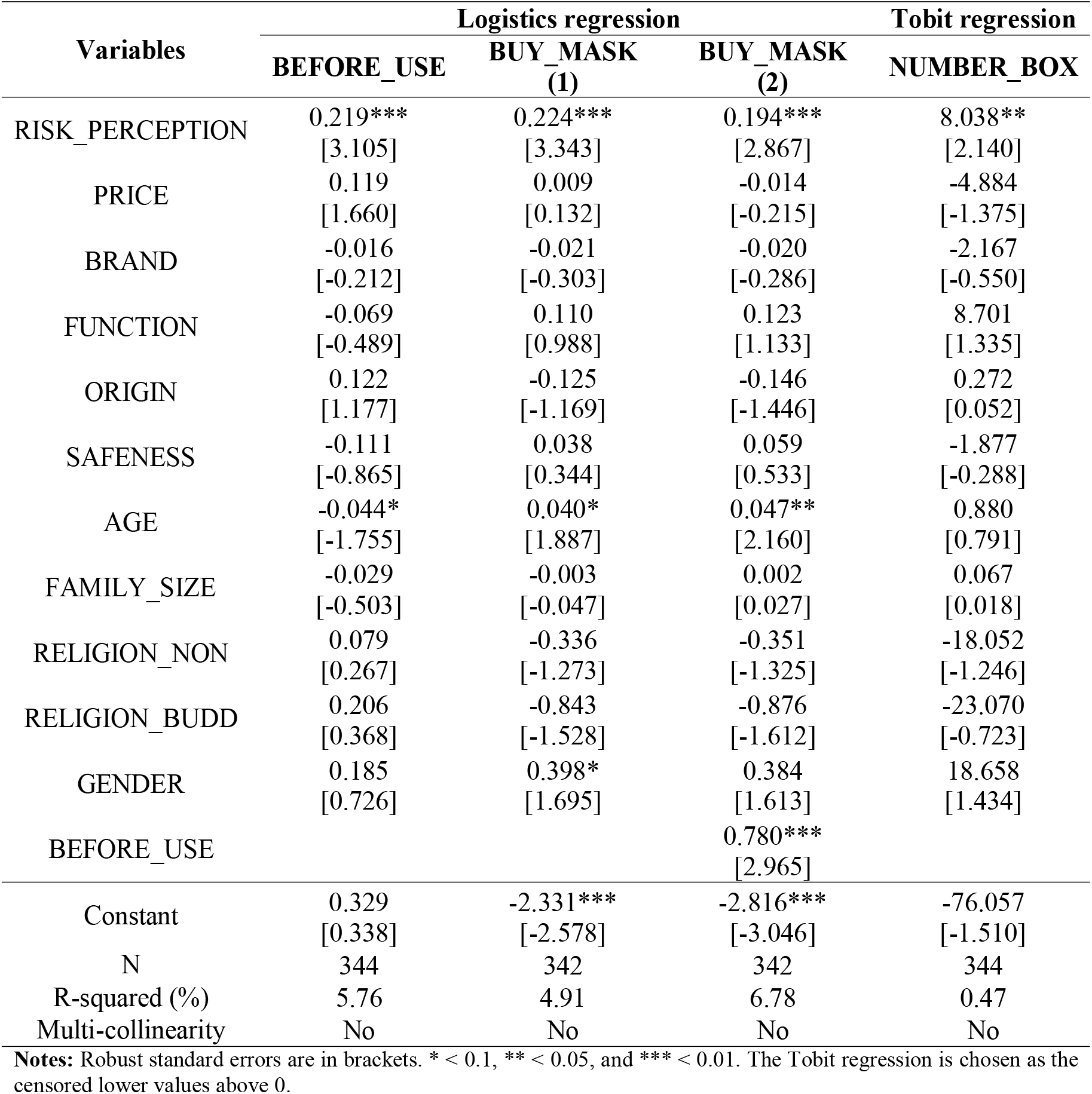
Regression to predict the behavior in using medical masks.

It is worth noting that the risk perception positively contributes to the likelihood to buy medical masks before and after the COVID-19 outbreaks at 1% significance level. It implies that people who perceive the danger of this novel virus are cautious to protect themselves against the possible transmission. Our study also confirmed the theoretical foundations devised by Leppin and Aro (2009). It means that risk is basically related to cognitions and emotions. Therefore, people perceived risk and would behave in the dominant expectancy and value components. Consequently, protective behaviors will be made to meet their cognition and emotions. However, we also draw the noticeable point that WHO’s recommendations do not include the behavior in using medical mask in public due to the lack of evidence. However, until now, the Asian countries, which have the habit of using masks, prove the efficiency to contain the COVID-19 outbreaks such as China, South Korea, Vietnam and so forth. Our study contributes to the existing literature by the relationship between risk perception and behavior in using masks.

In addition, we observe an evidence that there is a difference in age between the before and after the COVID-19 outbreak. The ‘age’ coefficient (0.04) is positive at 10% significance level after the COVID-19 outbreak. However, when we control the behaviors which are done before the COVID-19 pandemic, the ‘age’ effect tends to be stronger (0.047) and this coefficient is at 5% significance level. In addition, the age factor is negative at 10% significance level before the spread of COVID-19. This finding suggests that the elder has a tendency to not care about the medical mask before the COVID-19 outbreak while this behavior seems to change afterwards. Especially, this changing effect is driven by the precedent behaviors. Obviously, our results refer to the changes in risk-taking behaviors. It means that people tend to perceive higher risk when getting old (Huynh, 2020). It is also in line with the study of Vroom and Pahl (1971) that there is significant negative relationship between age and risk-taking behavior.

### 3.3. Robustness check

As mentioned earlier, there might be a high correlation among five features of medical masks. Therefore, we performed the Factor Analysis to examine how many factor that we can comprise these features. Finally, the result suggests generating one factor named FEATURES for medical masks. We estimated the Table 3 once again by replacing five factors with FEATURES. Then, the results remain robust. The results are available upon request.

## 4. Conclusion

It is too early to evaluate the effects of COVID-19 pandemic on the different aspects in our society. However, the contagious COVID-19 novel virus need to be studied urgently, especially how to contain the spreading in our community. Drawing a sample of 345 Vietnamese citizens from 15 to 47 years old, we found that the risk perception is the main factor which drives the behaviors in wearing medical masks in the public. This substantially contributes to the behavioral economics as well as infectious disease epidemiology to understand and creating behavior change to cope with this deadly virus. In addition, age is also a factor should be carefully examined later because we found that the elder is likely to wear masks after the COVID-19 outbreaks, with the weak evidence. Finally, those who have good habit in wearing mask are consistent to their behaviors.

Based on our results, we would suggest two main policies at that time. The communication about the COVID-19 risk is very important to drive the human behaviors. Although the effects of wearing masks are controversial, the Asian countries seem to control the COVID-19 pandemic better because the citizens tend to perceive higher risk in the public. Then, they choose to protect themselves by avoiding the virus droplet via closely social communication. Our study limits that the study sample covers those who aged 15 to 47 years. Therefore, the most vulnerable people (over 80 years old) might have different risk perception. We leave this avenue for the future research.

## Data Availability

The data will be available upon request.

## Disclaimer

All authors contributed in their individual capacities and the views that are expressed in this commentary do not necessarily reflect the views of their respective organizations. This study is approved by the scientific committee by School of Banking, University of Economics Ho Chi Minh City (Vietnam) for ethics declaration.

